# Interrupted Time Series Design and Analyses in Health Policy Assessment

**DOI:** 10.1101/2024.08.01.24311280

**Authors:** Huan Jiang, Jürgen Rehm, Alexander Tran, Shannon Lange

**Author notes:** Corresponding Author: Huan Jiang, 33 Ursula Franklin Street, Room T420, Toronto, Ontario, Canada, M5S 2S1.

## Abstract

Interrupted time series design is a quasi-experimental study design commonly used to evaluate the impact of a particular intervention (e.g., a health policy implementation) on a specific outcome. Two of the most often recommended analytical approaches to interrupted time series analysis are autoregressive integrated moving average (ARIMA) and Generalized Additive Models (GAM). We conducted simulation tests to determine the performance differences between ARIMA and GAM methodology across different policy effect sizes, with or without seasonality, and with or without misspecification of policy variables. We found that ARIMA exhibited more consistent results under certain conditions, such as with different policy effect sizes, with or without seasonality, while GAM were more robust when the model was misspecified. Given these findings, the variation between the models underscores the need for careful model selection and validation in health policy studies.

## 1 Introduction

Interrupted time series (ITS) analysis assesses whether an intervention is associated with a shift in the trend of the outcome of interest. ITS involves the assessment of an event or policy intervention and its effect on the trend of a target variable. An ITS design often entails collecting data both before and after a significant event, which could include a health policy intervention, or some other meaningful event (e.g., adoption of a new screening technique, large sociocultural movement etc.) and determining if it was associated with a change in the incidence or rate of an outcome. ITS is considered one of the best quasi-experimental designs when randomized controlled trials (RCTs) are not feasible (Cook, Campbell, & Shadish, 2002; Lopez Bernal, Cummins, & Gasparrini, 2018). There are a range of effects that can describe the impact of the intervention in an ITS model. For example, there could be an immediate level change or change in the time trend of the outcomes following the policy intervention. At times, a policy intervention can have either an immediate or a lagged effect, or both. For example, when a policy change consists of a ban or other restriction on alcohol marketing (Manthey, Jacobsen, Klinger et al., 2024), most of the effects are expected to involve a lag-time and the effect may take years to be fully realized. In addition, those effects need to considered based on the sample size and related statistical power to meaningfully interpret the findings. These factors need to be taking into account when designing an ITS study.

Analyzing time series data involves a few unique challenges due to three primary features: autocorrelation, non-stationarity, and seasonality. Autocorrelation refers to the correlation between each observation and observations at previous time points, which are often dependent on one other. As well, time series data are often not stationary because the variances of the observations change with time and may exhibit an increasing or decreasing trend (Hyndman & Athanasopoulos, 2018). Seasonality refers to variation of a frequency with regular time intervals, such as month of the year or day of the week. Common seasonality effects in health time series data are weather conditions, weekend or holiday effects, and administrative process patterns. For instance, all-cause mortality rates are higher in the winter months (Hajat & Gasparrini, 2016; Wilkinson, Pattenden, Armstrong et al., 2004).

A few systematic reviews have looked at ITS design and statistical models. (Jandoc, Burden, Mamdani et al., 2015) gathered over 200 studies in drug utilization and found ITS designs were used increasingly but reporting standards varied. (Ramsay, Matowe, Grilli et al., 2003) reviewed 58 studies and demonstrated that ITS were often underpowered, analyzed inappropriately, and reported poorly based on their quality criteria. In their review of 115 ITS studies, (Hudson, Fielding, & Ramsay, 2019) found the most commonly (78%) used analysis method was segmented regression. A similar conclusion was drawn by (Turner, Karahalios, Forbes et al., 2020), who identified 200 ITS studies that evaluated public health interventions or exposures from PubMed. They concluded that pre-specification of the statistical models was important. Though segmented regression is the most common approach, it assumes the residuals are independent and is therefore not reliable in the presence of autocorrelation and seasonality. Two alternatives are Autoregressive Integrated Moving Average (ARIMA) models and Generalized Additive Models (GAM). ARIMA modelling is considered to be a more flexible tool to evaluate health interventions and model different types of impacts (Schaffer, Dobbins, & Pearson, 2021). Generalized Additive Models (GAMs), on the other hand, can incorporate complex random effects that are common in real ITS, without requiring pre-specification of the form of the non-linear relationship. However, few studies have compared the accuracy of these two methods. As such, more guidance on the conduct and reporting of ITS studies is required to improve the study design and analysis.

In a methodological paper by (Beard, Marsden, Brown et al., 2019), the authors identified a series of ITS designs appropriate for addiction research. Furthermore, they described how ITS was being used, what design characteristics should be considered, and how the data should be analyzed. As an extension of their work, in this paper, we will describe the importance of pre-specifying the shape of intervention effects, the rationale for the number and spacing of data points selected, and the theory behind ITS models and how they can be used to evaluate population-level interventions, such as the introduction of health policies. Most importantly, we compare the performance of ARIMA and GAM—two analytical approaches to ITS analysis—under different assumptions.

## 2 Time series Design

### 2.1 Sample size consideration

Given the complexity of ITS data, there is no formula available to determine the minimum sample size for a time series analysis. Most time series experts provide a few rules of thumb. For example, (Warner, 1998) suggest a minimum of 50 observations are required to conduct a time series analysis. However, those rules of thumb are oversimplified and overlook the underlying variability of the data. Sample size, or number of data points required, depends on the number of parameters to be estimated and the degree of randomness. Consider a time series with seasonality, a seasonal model takes up an extra of m-1 degrees of freedom where m is the seasonal period (e.g., m=12 for monthly data). Therefore, a short series might not contain enough data for testing purposes and can only be analyzed with very simple models with one or two parameters. Such models might fail to identify an effect due to a Type II error. Since the power of the estimated parameters positively relate to the numbers of data points (Krone, Albers, & Timmerman, 2017), it must be emphasized that more data are required to obtain enough statistical power to detect the impact of a policy intervention, for example.

It is particularly challenging to conduct a proper power analysis for a GAM, since different smooth terms can have different effective degrees of freedom and different types of basic functions, depending on the nature of the relationship. To conduct a test of power for these complex models, a simulation approach that uses multiple sets artificial data to fit against an outcome may help to inform the necessary sample size required for a GAM.

### 2.2 Pre-specification of the shape of the intervention effect

It is also important to consider that effects might occur within a range around the implementation of the intervention. For example, when it was announced that an increase in taxation would be placed on alcohol in Lithuania, consumption of alcohol started declining in anticipation of this change (Rehm, Štelemėkas, Ferreira-Borges et al., 2021a). In other cases, the impact may be delayed by one or more-time units due to the course of a disease (e.g., liver cirrhosis, see (Skog, 1984; Tran, Jiang, Lange et al., 2022)). Policy interventions might have an immediate effect, a lagged effect or both. For example, alcohol prices or taxes might have an immediate effect on traffic fatalities, while having both immediate and lagged effects on liver cirrhosis mortality (Holmes, Meier, Booth et al., 2012).

We recommend pre-specifying a reasonable period of time in which it would be expected for the impact to be observed based on content knowledge or previous research to avoid spurious associations. The most appropriate lead or lag time within the range of options should be determined at the modelling stage.

## 3 Statistical method

Since repeated observations are often dependent sequentially, classic techniques like ITS must partition noise from real effect, which require more advanced statistical modelling. A variety of statistical models have been applied to examine the policy intervention effects with an ITS design. The two statistical models that have been less used, but are considered to have potential to be more accurate with present of autocorrelation, are the ARIMA and GAM. Each has distinct features, strengths, and weaknesses when applied.

### 3.1 ARIMA model

An ARIMA model (Rehm, Štelemėkas, Ferreira-Borges et al., 2021b) assumes that time series are stationary and invertible. Stationarity suggests a constant mean, variance, and autocorrelation over time after differencing. Invertibility implies that the model errors can be explained by the current and past forecast errors. The three main components for an ARIMA model are Autoregressive (AR), Integrated (I), and Moving Average (MA). The AR component represents the relationship between an observation and its previous (lagged) observations. Integrated component represents the number of differences needed to transform the time series into a stationary series. The Moving Average component captures the relationship between an observation and the residual errors obtained from previous forecasts.

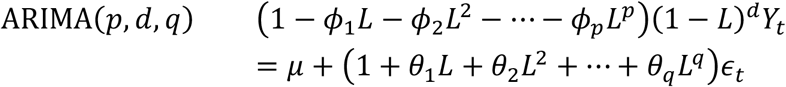

*Y*_*t*_ is the time series value at time t. *ϵ*_*t*_ is white noise. *ϕ*_*i*_ represents the parameters of the AR component. *θ*_*i*_ represents the parameters of MA component. *L* is the lag operator. So, *LY*_*t*_ would represent *Y*_*t*-1_, which is the value of the series lagged by one period. This is similar for *L*^2^ to *L*^*p*^. The left-hand side represents the autoregressive component of order *P* and the differencing of order d. The right-hand side includes the constant *μ* and represents the moving average component of order *q*.

The ARIMA model requires that the data are stationary, which is characterized by a stable mean and variance over time. It’s also crucial to ensure the residuals satisfy stationarity— that is, a mean of zero, constant variance, and no autocorrelation. Despite the ability of the ARIMA model to accommodate multiple predictors, its application is restricted to linear relationships with the dependent variable. The model particularly excels in short-term forecasting, as its predictions are heavily dependent on the recent past values and errors.

### 3.2 GAM model

GAM (Wood, Pya, & Säfken, 2016) is a type of regression model used to capture a nonlinear relationship between predictors and response variables without having to specify the form of the relationships. It also has few assumptions such as independence of observations, correct link function and variance function, and smoothness and additivity of the effect of predictors. Conversely, it offers a flexible approach to forecasting time series data. Its ability to capture more complex patterns makes it a preferable choice for certain datasets, especially since it can handle a non-constant variance of residuals. A significant advantage of GAM is its capability to analyze non-linear relationships while adjusting for potential covariates. Nevertheless, GAMs require that errors between observations are independent, a condition which is challenging when using time series data, where autocorrelation is a common feature.

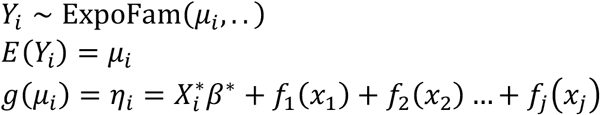

where the response variable *Y*_*i*_ follows an exponential family distribution. *g* is a monotonic link function. 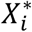 is the ith row of the model matrix of the parametric part of the model. *f*_*j*_ are the smooth functions of the covariates *x*_*j*_.

## 4 Simulation Study

Since ARIMA requires a stationary series and GAMs do not, they should be carefully selected for different types of data and different intervention effects. When comparing the results from the GAM and ARIMA models using actual policy intervention data, one will notice differences in the coefficient estimates of policy effects on the outcomes. To reveal the robustness and the differences of the two methods, a simulation study was employed to compare the ARIMA and GAM models using different assumptions.

We aimed to compare outcomes across different policy effect sizes, with or without seasonality, and with or without misspecification of policy variable. Simulated data was designed to mimic real-world scenarios spanning over a different length of time periods, incorporating a given ARIMA process, seasonality, and a 2-year delayed effect of a policy intervention. Subsequently, 100 datasets were simulated and analyzed using the respective ARIMA and GAM models with estimates such as coverage probability, average length of confidence interval (CI), and type II error rate. Before drawing any comparisons, the models were examined to ensure they met the stationary assumption. Ultimately, these models were assessed based on their precision in estimating the effect size of specific policy interventions.

The simulated outcomes were the sum of intercept, linear trend of time, policy impact, seasonal effects, ARIMA process, and random errors. The intercept was arbitrarily set as 100. There are five different policy effect sizes being investigated: 2, 5, 10, 15 and 20, e.g. the 2 intercept indicates that the policy reduces the mean outcome by 2. The policy variable was coded as an exponential distribution with a rate of 1 or 5, as recommended by experts. To replicate previous policy-intervention studies, we assumed the outcome would decrease by 0.3 each month, the outcomes contained an ARIMA (1,1) with the two coefficients being 0.6 and -0.8, and the random errors distributed with the mean being 12 and the standard error being 5.

### 4.1 Sample size consideration

Even when the sample size appears to be large enough, the power to detect an effect depends on when in the time series the intervention occurs. We simulated a study that contains 216 time points (months), where a policy intervention was introduced at the 60th, 108th, and 156th time points. When the modeled policy was assumed to have a lagged effect, and the time series was analyzed using the matched models, the coverage probability, the percentage of the 100 CIs that encompass the true value, ranged between 85.6%–90.7% (see results in Appendix), with the intervention introduced at the 108th time point, which has the highest coverage probability most of the scenarios. Even when the models were misspecified, the effect of intervention introduced at the 60th time point had a higher chance of being captured. That is, enough time points are present after the intervention implementation to correctly measure the intervention effect. We found that GAMs need more data points than ARIMA models with the monthly data due to the fact that estimates of spline functions are more complex than linear functions. Most time series experts suggest that at least 50 observations were required for the use of time series analysis (Warner, 1998), but in order to use GAMs, at least 100 observations were required in our simulation study.

### 4.2 Estimate intervention effect under various assumptions

We simulated a 10-year time series of monthly data that contained 120 observations with an intervention occurring at the 60th month. The simulated dataset was then analyzed using both ARIMA and GAM models. Tables 1 and 2 compare the ARIMA and GAM models across different policy effect sizes (the mean outcome being reduced by 2, 5, 10, 15, or 20) with or without seasonal effects, while the simulated data were analysed using matching statistical methods. Both models have a high likelihood of type II errors, indicating the frequency with which a model overlooks the genuine effect sizes, when the true effect size was less than 5. This implies that for smaller true effects within this range, both models struggle to discern the true effect from zero. There were notable differences when the true effect size was greater than 5: both models exhibited near zero type II error rate. That is, when models were correctly specified, with respect to seasonality, and the policy effects were significant enough, both ARIMA and GAM were able to identify the true effect. However, when the data did not exhibit seasonality, GAMs appeared to produce wider 95% CIs and higher Type II errors than ARIMAs.

**Table 1.**
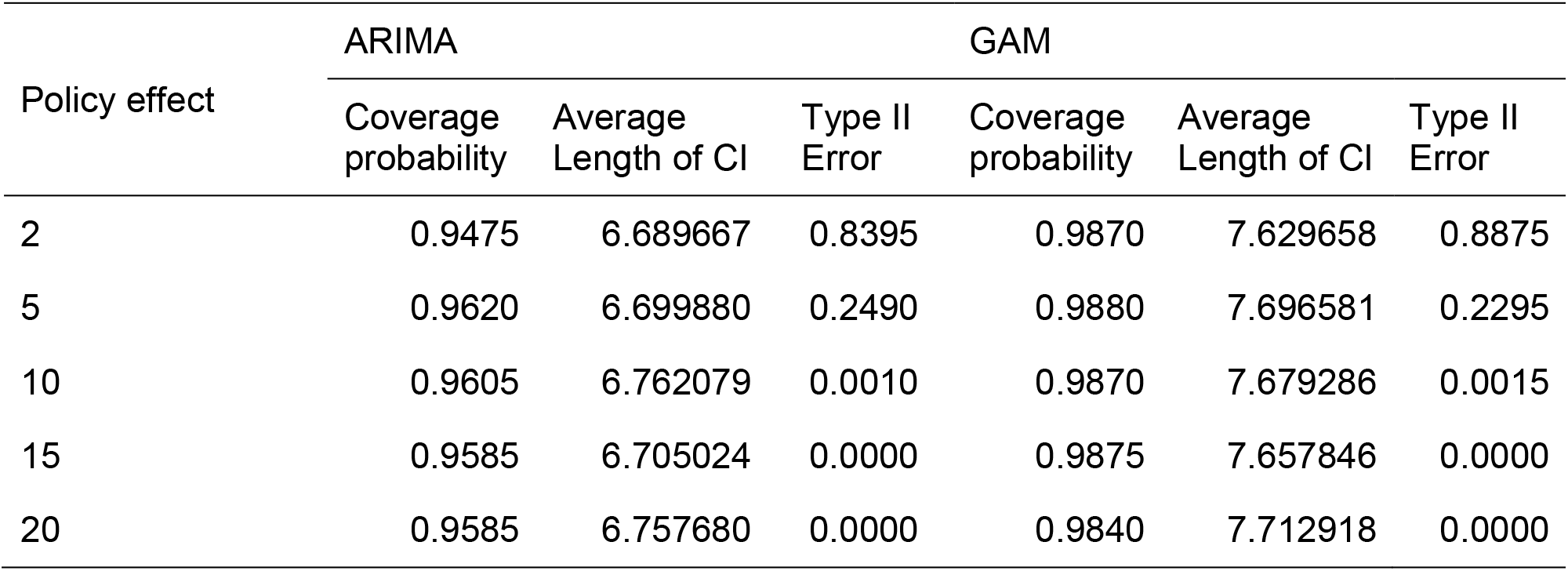
Coverage probability, average length of CI and Type II error rates for ARIMA and GAM, with seasonality, by policy effect size.

| Policy effect | ARIMA |  |  | GAM |  |  |
| --- | --- | --- | --- | --- | --- | --- |
|  | Coverage probability | Average Length of CI | Type II Error | Coverage probability | Average Length of CI | Type II Error |
| 2 | 0.9475 | 6.689667 | 0.8395 | 0.9870 | 7.629658 | 0.8875 |
| 5 | 0.9620 | 6.699880 | 0.2490 | 0.9880 | 7.696581 | 0.2295 |
| 10 | 0.9605 | 6.762079 | 0.0010 | 0.9870 | 7.679286 | 0.0015 |
| 15 | 0.9585 | 6.705024 | 0.0000 | 0.9875 | 7.657846 | 0.0000 |
| 20 | 0.9585 | 6.757680 | 0.0000 | 0.9840 | 7.712918 | 0.0000 |

**Table 2.** Coverage probability, average length of CI and Type II error rates for ARIMA and GAM, without seasonality, by policy effect size.

| Policy effect | ARIMA |  |  | GAM |  |  |
| --- | --- | --- | --- | --- | --- | --- |
|  | Coverage probability | Average Length of CI | Type II Error | Coverage probability | Average Length of CI | Type II Error |
| 2 | 0.9350 | 10.80147 | 0.8690 | 0.9700 | 13.90594 | 0.9330 |
| 5 | 0.9295 | 10.82204 | 0.5495 | 0.9710 | 13.96299 | 0.7415 |
| 10 | 0.9340 | 10.78308 | 0.0735 | 0.9740 | 13.95743 | 0.1670 |
| 15 | 0.9355 | 10.73325 | 0.0040 | 0.9715 | 13.87774 | 0.0155 |
| 20 | 0.9255 | 10.69711 | 0.0000 | 0.9585 | 13.87514 | 0.0000 |

### 4.3 Estimate intervention effect when models are misspecified

In Tables 3 and 4, we assessed misspecified models to determine their performance under suboptimal conditions. Here, we operate under the assumption that the policy effect in the model is inaccurately represented—coded as a dummy variable—whereas the actual data suggests the policy effect follows an exponential decay. In the first scenario, the policy variable was assigned the value of 1 for 24 months after policy implementation and 0 for the rest of the study time. In the second scenario, a common practice was used of assigning a value of 0 before the policy was introduced and a value of 1 thereafter. In both of those misspecified models, Type II error rates were higher with smaller effect sizes. However, in the second scenario, the ARIMA resulted in a high Type II error rate of 0.97 when the policy effect was 15, and one of over 0.50 when the policy effect was 20, while GAM had Type II errors of 0.21 and 0, respectively. Both scenarios produced a wider CI when compared to models without misspecification.

**Table 3.** Coverage probability, average length of CI and Type II error rates for ARIMA and GAM, with policy misspecification, by policy effect size, where policy variable was assigned the value of 1 for 24 months after policy implement and 0 for the rest of the study time

| Policy Effect | ARIMA<br>CP | ARIMA CI<br>AL | Gam<br>CP | Gam CI<br>AL | ARIMA<br>Type II Error | Gam<br>Type II Error |
| --- | --- | --- | --- | --- | --- | --- |
| 2 | 0.9655 | 13.08090 | 0.9995 | 15.64162 | 0.9195 | 0.9925 |
| 5 | 0.9700 | 14.17441 | 0.9990 | 15.98781 | 0.7115 | 0.8305 |
| 10 | 0.9930 | 17.32117 | 0.9960 | 17.43877 | 0.2215 | 0.2055 |
| 15 | 0.9855 | 20.46541 | 0.9820 | 21.16744 | 0.0335 | 0.0585 |
| 20 | 0.9830 | 24.82339 | 0.9830 | 29.52140 | 0.0050 | 0.0945 |

\* CP and AL stands for coverage probability and average length, respectively

**Table 4.**
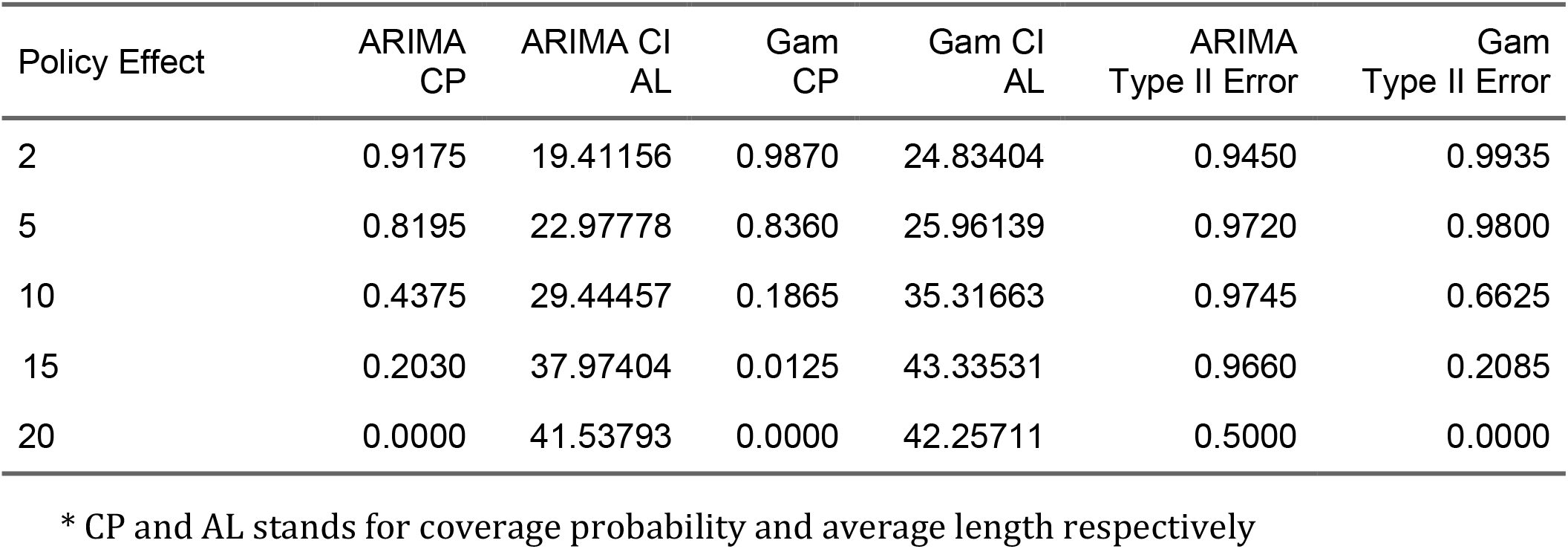
Coverage probability, average length of CI and type 2 error rates for ARIMA and GAM, with policy misspecification, by policy effect size, where the policy variable was assigned a value of 0 before policy being introduced and 1 after.

**Table 5.** Coverage probability, average length of CI and Type II error rates for ARIMA and GAM, with outliers in the outcome, by policy effect size.

| Policy Effect | ARIMA<br>CP | ARIMA CI<br>AL | Gam<br>CP | Gam CI<br>AL | ARIMA<br>Type II Error | Gam<br>Type II Error |
| --- | --- | --- | --- | --- | --- | --- |
| 2 | 0.9370 | 22.22706 | 0.9715 | 24.97442 | 0.9245 | 0.9625 |
| 5 | 0.9330 | 22.58963 | 0.9755 | 25.39765 | 0.8365 | 0.8935 |
| 10 | 0.9585 | 25.00787 | 0.9705 | 28.10154 | 0.6055 | 0.6560 |
| 15 | 0.9670 | 28.25577 | 0.9695 | 32.66184 | 0.3450 | 0.4040 |
| 20 | 0.9585 | 31.95723 | 0.9585 | 37.47694 | 0.1880 | 0.2470 |

\* CP and AL stands for coverage probability and average length respectively

### 4.4 Estimates of intervention effects when data contain outliers

To assess how robust the two models are in the presence of outliers comprising 10% of the outcome, simulated outliers were generated to exceed either three times the standard deviations or fall below this threshold. When outliers were present, ARIMA and GAM performed similarly with type II errors decreasing when the policy effect increased. In addition, the CIs were wider than those without outliers.

## 5 Discussion

Accurate modeling and forecasting of the impact of policy interventions can inform policymakers about the efficacy of their interventions and guide future actions. Based on the findings here, we advocate for predetermining an appropriate timeframe for observing intervention effects as well as drawing from subject expertise or prior studies in advance in order to prevent drawing misleading conclusions. We compared GAM and ARIMA, two popular time series models, using simulated data with certain patterns or trends one would expect to see in the real world following a policy intervention, e.g., a policy with a lagged effect.

When ARIMA and GAM were applied to data that did not exhibit seasonality and was stationary, the CIs of estimates were much wider when using GAM. That is, the estimates were less precise when applied to outcomes that did not contain a seasonal pattern. Therefore, it is important to scrutinize the outcomes for seasonality before the use of any models. In cases where there are no repeating cycles over time and there is confidence in the accurate definition of an intervention policy, the ARIMA model offers more precise estimates with lower Type II error rates. Additionally, it outperforms GAM by providing estimates that are more precise and exhibit lower Type II error rates in situations where 10% of the outcome are outliers.

Another important finding is the robustness of GAM models when the policy effects were expected and coded inconsistently with the simulated condition. When the effect size was misspecified but the effect period of the policy intervention was correctly specified, both GAM and ARIMA demonstrated comparable performance. Conversely, when both the effect size and intervention period were misspecified, ARIMA models failed to detect any policy effect, even when the policy significantly reduced the outcome by 15%. In contrast, GAM models excelled in correctly identifying the effects of policy intervention. This implies that the GAM model should be preferred over the ARIMA model when the impact of a policy intervention is unknown or uncertain.

While our simulation study provides some valuable insights into model performance under controlled conditions, caution must be exercised when extrapolating these results to real-world scenarios since they are more complex than simulated data. Further, future research should expand the simulated data to incorporate multiple policies and their interactions. Such an approach would offer a deeper understanding, especially since real-world settings often entail the simultaneous implementation of multiple policies.

## 6 Conclusion

In conclusion, when using ITS modelling, we suggest specifying a reasonable timeframe at the design stage, within which the expected policy impact should be observed. Both ARIMA and GAM models provide insights into the investigation of policy effects. Nevertheless, their different performances in various simulated scenarios underscore the significance of thorough examination of outcomes, precise specification of policy variables, and careful selection and validation of the appropriate model.

## Data Availability

All data produced in the present study are available upon reasonable request to the authors

## Appendix

**Table A1:** Comparison of ARIMA and GAM models under different policy effects with simulated data containing 216 time points, where a policy intervention was introduced at the 60^th^ time point.

| Policy Effect | ARIMA<br>CP | ARIMA CI<br>AL | Gam<br>CP | Gam CI<br>AL | ARIMA<br>Type II Error | Gam<br>Type II Error |
| --- | --- | --- | --- | --- | --- | --- |
| 0.1 | 0.93 | 9.476391 | 0.97 | 13.29382 | 0.92 | 0.97 |
| 2 | 0.88 | 9.484434 | 0.95 | 13.34967 | 0.80 | 0.92 |
| 5 | 0.89 | 9.544705 | 0.97 | 13.35895 | 0.44 | 0.76 |
| 10 | 0.95 | 9.477075 | 0.97 | 13.15322 | 0.03 | 0.15 |
| 20 | 0.93 | 9.934477 | 0.98 | 13.56173 | 0.00 | 0.00 |

**Table A2:**
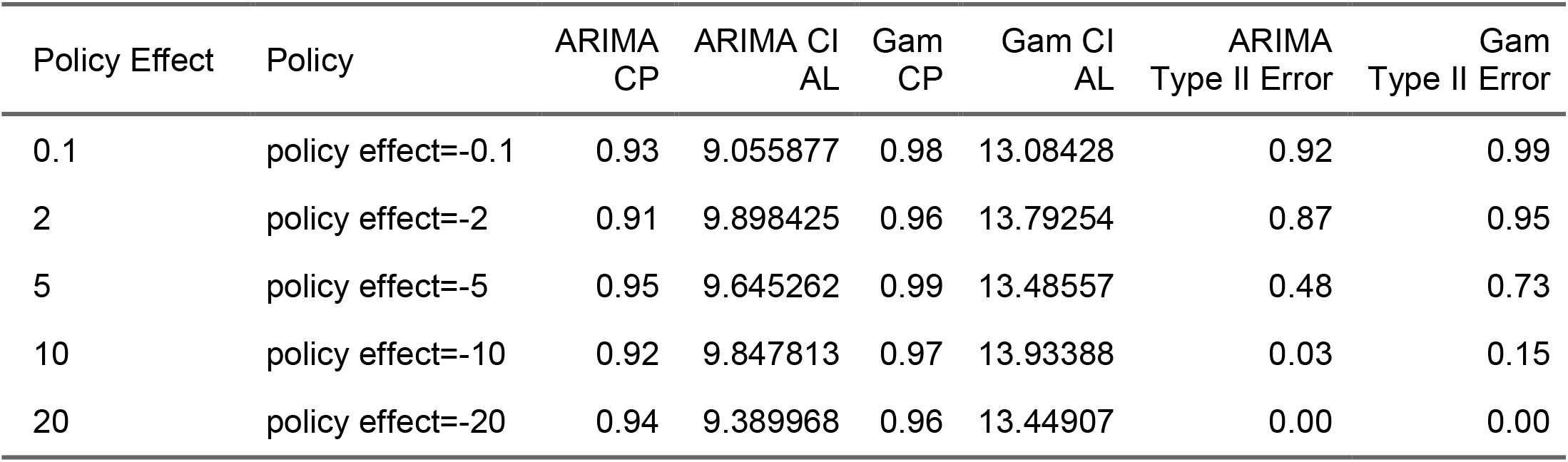
Comparison of ARIMA and GAM models under different policy effects with simulated data containing 216 time points, where a policy intervention was introduced at the 108^th^ time point.

| Policy Effect | Policy | ARIMA<br>CP | ARIMA CI<br>AL | Gam<br>CP | Gam CI<br>AL | ARIMA<br>Type II Error | Gam<br>Type II Error |
| --- | --- | --- | --- | --- | --- | --- | --- |
| 0.1 | policy effect=-0.1 | 0.93 | 9.055877 | 0.98 | 13.08428 | 0.92 | 0.99 |
| 2 | policy effect=-2 | 0.91 | 9.898425 | 0.96 | 13.79254 | 0.87 | 0.95 |
| 5 | policy effect=-5 | 0.95 | 9.645262 | 0.99 | 13.48557 | 0.48 | 0.73 |
| 10 | policy effect=-10 | 0.92 | 9.847813 | 0.97 | 13.93388 | 0.03 | 0.15 |
| 20 | policy effect=-20 | 0.94 | 9.389968 | 0.96 | 13.44907 | 0.00 | 0.00 |

**Table A3:** Comparison of ARIMA and GAM models under different policy effects with simulated data containing 216 time points, where a policy intervention was introduced at the 156^th^ time point.

| Policy Effect | ARIMA<br>CP | ARIMA CI<br>AL | Gam<br>CP | Gam CI<br>AL | ARIMA<br>Type II Error | Gam<br>Type II Error |
| --- | --- | --- | --- | --- | --- | --- |
| 0.1 | 0.904 | 13.05055 | 0.999 | 23.73328 | 0.905 | 0.999 |
| 2 | 0.907 | 13.56236 | 0.999 | 23.76087 | 0.869 | 0.997 |
| 5 | 0.949 | 17.98891 | 0.988 | 24.23383 | 0.849 | 0.996 |
| 10 | 0.988 | 26.10929 | 0.901 | 26.64836 | 0.916 | 0.929 |
| 20 | 0.917 | 38.49901 | 0.086 | 41.77918 | 0.879 | 0.558 |

### Optimization procedure of ARIMA model

In order to perform the optimization procedure such as the maximum likelihood estimation to determine the best ARIMA model parameters, the ARIMA model would need to transform into its state space and Markovian state space form, which involves representing the ARIMA model as a system of states that evolve over time, where each state only depends on the previous state. For the ARIMA model in its state-space representation, the state refers to the unobservable variables, and the observations are the values in the time series. It would start with initializing state estimate *Z*(*t* + 1 | *t*) and state covariance *P*(*t* + 1 | *t*) = *P*(0 | 0). Then we would predict the next state and its covariance using the state transition model. Next would be to calculate the Kalman Gain to determine the weight given to the prediction error when updating the state estimate. Assuming the errors are Gaussian (a common assumption for ARIMA models), the log likelihood for the entire observations is just the sum of the log-likelihoods from each step. The function is then optimized to estimate the best-fit parameters of the ARIMA model.

### Optimization procedure of GAM model

Penalized Iteratively Reweighted Least Squares is the iterative algorithm used to estimate the parameters of the model, when a penalty is applied to some parameters for smoothness. With the initial working response and the weights (diagonal weight matrix from the link function’s derivative), and the penalty matrix, one can solve for the penalized weighted least squares and obtain the new estimated *α* and the smooth function *f*_*j*_(*x*_*j*_). Thus the updated *η* can be calculated *η* = *Xβ* + Σ_*i*_ Σ_*k*_ *b*_*k*_ (*x*)*α*_*k*_ and *μ*, and the expected value of the response can then be obtained from *μ* = *g*^-1^(*η*). After assessing whether the parameters have converged, by evaluating the change in some criterions like deviances, the algorithm either returns to the initiation step with the new values of *α* and *μ* to refine the next round of estimates or stops entirely.

## Acknowledgements

The authors would like to acknowledge Astrid Otto for help with copy-editing the final version of this manuscript as well as the National Institute on Alcohol Abuse and Alcoholism (NIAAA) (Award Number 1R01AA028224) of the National Institutes of Health for funding this research.

## Author Statement

HJ led the conception and design of the study, conducted the statistical analyses and contributed to the writing of the manuscript. JR, SL, AT assisted with the interpretation of results and revised the intellectual content of the manuscript. All authors approved the final version of this manuscript.

